# Plasmatic HIV-1 soluble gp120 is associated with immune dysfunction and inflammation in ART-treated individuals with undetectable viremia

**DOI:** 10.1101/2023.08.15.23294128

**Authors:** Mehdi Benlarbi, Jonathan Richard, Catherine Bourassa, William D. Tolbert, Carl Chartrand-Lefebvre, Gabrielle Gendron-Lepage, Mohamed Sylla, Mohamed El-Far, Marc Messier-Peet, Camille Guertin, Isabelle Turcotte, Rémi Fromentin, Myriam Maude Verly, Jérémie Prévost, Andrew Clark, Walther Mothes, Daniel E. Kaufmann, Frank Maldarelli, Nicolas Chomont, Philippe Bégin, Cécile Tremblay, Jean-Guy Baril, Benoit Trottier, Sylvie Trottier, Ralf Duerr, Marzena Pazgier, Madeleine Durand, Andrés Finzi, the Canadian HIV, Aging Cohort Study

**Affiliations:** Centre de Recherche du CHUM, Montréal, QC, Canada; Département de Microbiologie, Infectiologie et Immunologie, Université de Montréal, Montreal, QC, Canada; Infectious Disease Division, Department of Medicine, Uniformed Services University of the Health Sciences, Bethesda, MD, USA; Department of Radiology, Radiation Oncology and Nuclear Medicine, Université de Montréal, Montreal, QC, Canada; McGill University, Montreal, QC, Canada; ViiV Healthcare, Global Medical Affairs, Middlesex, United Kingdom; Department of Microbial Pathogenesis, Yale University School of Medicine, New Haven, CT, USA; Division of Infectious Diseases, Department of Medicine, Lausanne University Hospital and University of Lausanne, Lausanne, Switzerland; HIV Dynamics and Replication Program, NCI, NIH, Bethesda, MD 20892, USA; Section of Allergy, Immunology and Rheumatology, Department of Pediatrics, CHU Sainte-Justine, Montréal, QC, Canada; Department of Medicine, Faculty of Medecine, Centre Hospitalier de l’Université de Montréal, Montréal, QC, Canada; Clinique de médecine urbaine du Quartier latin; Département de médecine familiale, Université de Montréal; Département de microbiologie, Université Laval; Vaccine Center, NYU Grossman School of Medicine, New York, NY 10016, USA; Department of Medicine, NYU Grossman School of Medicine, New York, NY 10016, USA; Department of Microbiology, NYU Grossman School of Medicine, New York, NY 10016, USA

**Keywords:** HIV-1, soluble gp120, anti-cluster A antibodies, chronic inflammation, cardiovascular diseases, atherosclerosis, immune dysfunction

## Abstract

**Background:** Chronic inflammation persists in some people living with HIV (PLWH), even during antiretroviral therapy (ART) and is associated with premature aging. The gp120 subunit of the HIV-1 envelope glycoprotein can shed from viral and cellular membranes and can be detected in plasma and tissues, showing immunomodulatory properties even in the absence of detectable viremia. We evaluated whether plasmatic soluble gp120 (sgp120) and a family of gp120-specific anti-cluster A antibodies, which were previously linked to CD4 depletion *in vitro*, could contribute to chronic inflammation, immune dysfunction, and sub-clinical cardiovascular disease in participants of the Canadian HIV and Aging cohort (CHACS) with undetectable viremia.

**Methods:** Cross-sectional assessment of plasmatic sgp120 and anti-cluster A antibodies was performed in 386 individuals from CHACS. Their association with pro-inflammatory cytokines, as well as subclinical coronary artery disease measured by computed tomography coronary angiography was assessed using linear regression models.

**Results:** In individuals with high levels of sgp120, anti-cluster A antibodies inversely correlated with CD4 count (p=0.042) and CD4:CD8 ratio (p=0.004). The presence of sgp120 was associated with increased plasma levels of IL-6. In participants with detectable atherosclerotic plaque and detectable sgp120, sgp120 levels, anti-cluster A antibodies and their combination correlated positively with the total volume of atherosclerotic plaques (p=0.01, 0.018 and 0.006, respectively).

**Conclusion:** Soluble gp120 may act as a pan toxin causing immune dysfunction and sustained inflammation in a subset of PLWH, contributing to the development of premature comorbidities. Whether drugs targeting sgp120 could mitigate HIV-associated comorbidities in PLWH with suppressed viremia warrants further studies.

**Key points:** Soluble gp120 is detected in the plasma of people living with HIV-1 with undetectable viremia. The presence of soluble gp120 and anti-cluster A antibodies is associated with immune dysfunction, chronic inflammation, and sub-clinical cardiovascular disease.

## INTRODUCTION

While antiretroviral therapy (ART) efficiently inhibits viral replication, people living with HIV (PLWH) have a 15-year gap in comorbidity-free years [1]. The underlying causes for this gap are numerous and include the presence of residual immune dysfunction and chronic antigenic stimulation by HIV, which contributes to a state of sustained inflammation [2]. Persistent immune dysfunction has also been associated with immunological non-response that many PLWH experience [3–7]. These individuals, despite efficient viral control, fail to restore circulating CD4^+^ T cells and have persistent immune activation, possibly leading to increased comorbidities.

The HIV-1 envelope (Env) glycoprotein, in particular the gp120 domain of the gp120/gp41 heterodimer, besides its role in viral entry, has been shown to exhibit a variety of immunoregulatory activities that could contribute to HIV-1 pathogenesis and immune dysfunction. Due to its non-covalent interaction with gp41, gp120 can be spontaneously released from the surface of virions and infected cells [8–10]. Consequently, variable amounts of soluble gp120 (sgp120) can be detected in plasma and tissues from PLWH, even during ART [7, 11, 12]. sgp120 has been associated with HIV-1-induced immune dysfunction in several studies [4–7]. sgp120 has also been reported to exert proinflammatory activities, as binding of gp120 to CD4 on the surface of monocytes, macrophages, T cells and dendritic cells has been found to induce the production of cytokines, including interleukin (IL)-6, IL-10, IL-1β, interferon alpha (IFN-α), IFN-γ and tumor necrosis factor alpha (TNF-α) [4, 13–15].

Notably, gp120 shed from productively infected cells has been shown to interact with CD4 present on uninfected bystander CD4^+^ T cells [15, 16]. This interaction leads to exposure of CD4 induced (CD4i) Env epitopes and sensitization of uninfected bystander CD4^+^ T cells to Antibody Dependent Cellular Cytotoxicity (ADCC) mediated by HIV+ plasma [16, 17]. Within HIV+ plasma, antibodies targeting conserved CD4i gp120 cluster A epitopes have been shown to elicit potent ADCC activity against sgp120-coated cells [15, 18]. Antibodies recognizing the gp120 inner domain cluster A region have been found to be responsible for most of the ADCC activity exhibited by chronically HIV-1-infected individuals, provided that the epitopes recognized by them are exposed [19]. Altogether, these observations suggest that soluble gp120 is associated with chronic inflammation, causing persistent antigen stimulation even in the context of prolonged ART and undetectable viremia in plasma, and a potential contributing factor of increased comorbidity risk. Here, we investigated whether the presence of sgp120 and ADCC-mediating antibodies, specifically anti-cluster A antibodies, were associated with immune dysfunction, chronic inflammation, and sub-clinical cardiovascular disease. To this end, we optimized an ELISA based assay to measure sgp120 in the plasma of 386 long-term treated PLWH with undetectable viremia from the Canadian HIV and Aging Cohort Study (CHACS) [20].

## METHODS

### Ethics statement

This research project was reviewed and approved by the CRCHUM Research Ethics Board (Ethics Committee approval number 22.173, 11.063, and 11.062.). Written informed consent was obtained from all study participants, and the research adhered to the standards indicated by the Declaration of Helsinki.

### Study design

We designed a cross-sectional study, nested within the CHACS. The specific objectives of the study were to quantify sgp120 in plasma of participants and study the relationship of sgp120 and anti-cluster A antibodies on immune dysfunction, inflammation, and sub-clinical cardiovascular disease.

### Study population

The CHACS protocol has been described previously [20]. Briefly, it is a prospective cohort study and biobank, ongoing since 2012 in 10 clinical sites across Canada. Inclusion criteria are to be 40 years old or older, or to have lived with HIV for at least 15 years and have a life expectancy of more than 1 year at enrollment. Participants with no known cardiovascular disease and a 10-year Framingham risk score of overt cardiovascular disease ranging from 5 to 20%, no allergy to contrast media, and no renal failure, were offered to participate to the cardiovascular imaging sub-study, by which they underwent computed tomography coronary angiography (CCTA – methods described below). Participants were selected for the present study if they were recruited into the Canadian HIV and Aging cohort study Montréal and Québec CHACS study sites and had undetectable viral load. For all participants, data on socio-demographic characteristics, HIV disease history, as well as traditional cardiovascular risk factors are available through the CHACS study database.

### Coronary Computed Tomography Angiography (CCTA)

CCTA was performed with a 256-section CT scanner (Brilliance iCT; Philips Healthcare) and 370 mg/mL iopamidol (Bracco Imaging) at a rate of 5 mL/sec with prospective electrocardiographic gating. Gantry rotation time was 270 msec, axial section thickness was 0.625 mm, and section interval was 0.45 mm. Plaques were identified on the CT angiograms by one board-certified radiologist (C.C.L., with 17 years of experience in cardiac CT) and plaque quantification was then performed by a research assistant (I.B., with 4 years of experience in plaque volume analysis). Plaque volume was assessed by using a semiautomated software (Aquarius iNtuition 4.4.6; TeraRecon) [21]. Analysis was performed in multiplanar reformat. Proximal and distal plaque boundaries were established visually. Segmentation was made by using the software with manual adjustment when plaque delimitation was inaccurate. Volume of plaque is expressed as total plaque volume (TPV), representing the volume of all plaques per participant. Image assessors were blinded to HIV status and cardiovascular risk factors. We previously established an excellent inter- and intraobserver agreement for plaque volume with this technique (intraclass correlation coefficient, 0.95 and 0.93, respectively) [22]. Results of plaque measurement in the CHACS have been reported previously [23].

### Enzyme-linked immunosorbent Assay

The sandwich ELISA was described elsewhere, with modifications [7, 15]. For measurement of soluble gp120, the gp120 N-terminus recognizing C11 monoclonal antibody (mAb) was coated (4 µg/mL in PBS) in parallel with bovine serum albumin (4 µg/mL in PBS) as a negative control. After blocking, diluted plasma 1:100 from HIV-infected or uninfected individuals were added to the well, followed by the addition of the CD4-binding site (CD4BS) specific broadly neutralizing HRP-conjugated N6 mAb (6 µg/mL) to the wells. HRP enzyme activity was determined after the addition of a 1:1 mix of Western Lightning oxidizing and luminol reagents (Perkin Elmer Life Sciences, Waltham, MA, USA). Light emission was measured with an LB942 Tri-Star luminometer (Berthold Technologies, Bad Wildbad, Germany). Signal obtained with BSA was subtracted for each plasma. A standard curve was established using two-fold dilutions (from 5ng to 10pg) of monomeric soluble HIV-1_YU2_ gp120 spiked into plasma from uninfected donors and used to normalize the signal obtained among different experiments. The positivity threshold was established using the following formula: mean of 20 plasma from uninfected donors + (3 standard deviation of the mean of the 20 plasma from uninfected donors). High levels of sgp120 were arbitrarily set as having sgp120 levels above the mean of all 20 plasma from uninfected donors + (6 standard deviation of the mean of all 20 plasma from uninfected donors).

For measurement of anti-cluster A antibodies, wells were coated with stabilized gp120 inner domain ID2 [24] (0.1 µg/mL in PBS), in parallel with BSA (0.1 µg/mL in PBS). After blocking, the cluster A specific A32 mAb (1 µg/mL) or diluted plasma 1:1000 from HIV-infected or uninfected individuals were added to the well and detection of plasma antibodies was performed using HRP-conjugated goat-anti-human IgG (Invitrogen) at a dilution of 1:3000. Signal was measured as described above and signal obtained with BSA was subtracted for each plasma and were then normalized to the signal obtained with A32 mAb present in each plate.

### Protein production and purification

Production and purification of monomeric soluble HIV-1_YU2_ gp120 was described elsewhere [15, 25]. Briefly, recombinant HIV-1_YU2_ gp120 was produced using a plasmid (pcDNA3.1) encoding the codon-optimized full-length HIV-1_YU2_gp120 containing a C-terminal hexa-histidine tag [9]. FreeStyle 293F cells (Thermo Fisher Scientific) were grown in FreeStyle 293F medium (Thermo Fisher Scientific) to a density of 1 × 10^6^ cells/mL at 37°C with 8% CO2 with regular agitation (150 rpm). Cells were transfected with the gp120 expressor using ExpiFectamine 293 transfection reagent, as directed by the manufacturer (Thermo Fisher Scientific). One week later, cells were pelleted, and supernatants were filtered using a 0.22-μm-pore-size filter (Thermo Fisher Scientific). Recombinant gp120 was purified by nickel affinity columns, as directed by the manufacturer (Thermo Fisher Scientific). Monomeric gp120 was subsequently purified by fast protein liquid chromatography (FPLC), as previously reported [25]. The purification by FPLC was performed using an ÄKTAprime Plus FPLC with a HiLoad 16/60 Superdex 200 PG (GE Healthcare, Chicago, IL, USA). The gp120 preparations were dialyzed against phosphate-buffered saline (PBS) and stored in aliquots at −80°C until further use. To assess purity, recombinant proteins were loaded on non-reducing SDS-PAGE polyacrylamide gels and stained with Coomassie blue.

### Multiplex measurements of soluble inflammatory markers

Duplicates of HIV-1 inactivated plasma samples were analyzed using a customized Human Magnetic Luminex Assay (LXSAHM-14, LXSAHM-1 or LXSAHM-2, R&D Systems; see Table S4 for complete analyte list). Plates were read using a MAGPIX system (Luminex) and analyzed with the software xPONENT v.4.3.229.0. Plasma samples presenting markers below the limit of detection were adjusted to 0. Results were adjusted according to sample dilution to represent the concentration in pg/mL of plasma.

### Measurement of HIV DNA

HIV DNA was measured as described previously [26]. Briefly, CD4^+^ T cells were isolated from PBMCs by negative selection using the EasySep Human CD4^+^ T cell Enrichment Kit (StemCell). HIV DNA was co-extracted using the AllPrep DNA/RNA Mini Kit (Qiagen). HIV DNA (LTR-gag) copies were measured using ultrasensitive nested PCRs [27]. Results were expressed as HIV DNA copies per million CD4^+^ T cells.

### Statistical Analysis

Appropriate descriptive statistics were used for normally and non-normally distributed variables, and variables were log-transformed, as needed. Differences in baseline characteristics between the participants included into subgroups and the remainder of the samples were assessed using Fisher’s exact and k-median tests for categorial and continuous variables, respectively. We hypothesized a priori that anti-cluster A antibodies and levels of sgp120 would interact to cause immune dysfunction and organ damage. Therefore, the associations between anti-cluster A antibodies and CD4 levels, CD4:CD8 ratios were modelled using linear regression models, including levels of sgp120 modelled as categorical (undetectable, low or high) and an interaction term between anti-cluster A antibodies and sgp120 levels. The association between the levels of sgp120, anti-cluster A antibodies and cardiovascular disease was modelled using logistic regression for the outcome of presence vs absence of coronary artery plaque (TPV=0 v. TPV>0). In participants with subclinical cardiovascular disease (TPV>0) and detectable levels of sgp120, logistic regression was performed for association between the levels of sgp120, anti-cluster A antibodies and the combination of sgp120 and anti-cluster A antibodies with the total volume of coronary atherosclerotic plaque. All models were adjusted for potential confounders (identified a priori based on clinical knowledge) using a parsimonious approach: potential confounders were kept into the models if they modified the point estimate for the main association by more than 10%. Absence of multicollinearity was ensured by examination of correlation matrices. Missing data for covariates was simulated by multiple imputations by chained equations. Correlations between multiplex biomarkers were calculated using Spearman’s Rank test and graphed as circular network in undirected mode using ggraph, igraph, and RColorBrewer packages in program R (R Core Team, 2014). Statistical analysis was conducted using Graphpad Prism version 8.4.2, Stata version 17 and R. For all analysis, alpha was set to 0.05. All hypothesis tests were two-sided, and no corrections were made for multiple testing.

## RESULTS

Plasma from 386 participants was available for measurements of sgp120 and anti-cluster A antibodies, 157 of whom also had measurements of soluble inflammatory markers by multiplex platform. Of the 386, 145 took part in the cardiovascular imaging sub-studies. The baseline characteristics of all participants and in each subgroup are summarized in Table 1. There were no differences in terms of socio-demographic characteristics, immune parameters, or traditional cardiovascular risk factors between the total study group and the two sub-groups.

**Table 1.** Characteristics of 386 participants to the Canadian HIV and Aging cohort study with undetectable HIV viral load, and participants to subgroups.

|  |  |  |  |  |  |
| --- | --- | --- | --- | --- | --- |
|  | Total study population | Cardiovascular imaging sub study | P value<sup>a</sup> | Multiplex sub-study | P value<sup>a</sup> |
| **Number** | 386 | 145 | -- | 157 | -- |
| **Age** (mean, SD) | 55.8 (7.6) | 56.6 (6.5) | 0.14 | 56.6 (6.6) | 0.07 |
| **Male Sex** (n, %) | 347 (90.0) | 134 (92.4) | 0.23 | 144 (91.7) | 0.39 |
| **Ethnicity** (n, %) |  |  |  |  |  |
| Caucasian | 317 (82.1) | 123 (84.8) | 0.39 | 131 (83.4) | 0.29 |
| Black | 32 (8.3) | 12 (8.2) |  | 15 (9.6) |  |
| other | 37 (9.6) | 10 (6.9) |  | 11 (7.0) |  |
| **Duration of HIV infection** (in years, median, IQR) | 18.3 (12-25) | 18.3 (12-25) | 0.27 | 19.0 (14-25) | 0.52 |
| **Nadir CD4** (median, IQR) | 210 (120-310) | 200 (90-300) | 0.30 | 195 (100-300) | 0.11 |
| **Less than 200** (n, %) | 155 (49.1) | 67 (52.8) |  | 74 (54.4%) |  |
| **Absolute CD4** (mean, SD) | 615 (250) | 620 (255) | 0.27 | 617 (251) | 0.50 |
| **Absolute CD8** (mean, SD) | 763 (376) | 796 (382) | 0.10 | 809 (408) | 0.07 |
| **CD4:CD8 ratio** (mean, SD) | 0.96 (0.53) | 0.96 (0.47) | 0.92 | 0.93 (0.49) | 0.35 |
| **Duration of ARV** (mean, SD) | 13.9 (6.7) | 14.1 (6.8) | 0.58 | 14.1 (6.9) | 0.58 |
| **Smoking status** (n,%) |  |  | 0.22 |  | 0.44 |
| Never | 128 (33.5) | 42 (29.4) |  | 48 (31.0) |  |
| Past or present | 254 (66.5) | 101 (70.6) |  | 107 (69.0) |  |
| **High blood pressure** (n, %) | 113 (29.7) | 45 (31.5) | 0.56 | 51 (32.7) | 0.31 |
| **Diabetes** (n, %) | 37 (9.6) | 15 (10.4) | 0.71 | 18 (11.5) | 0.30 |
| **Framingham risk score** (median, IQR) | 9 (5-16) | 9 (6-13) | 0.72 | 9 (6-13) | 0.58 |
| **Log anti-cluster A antibodies** (mean, SD) | 2.4 (1.4) | 2.2 (1.2) | 0.22 | 2.3 (1.2) | 0.71 |
| **sgp120** (n,%) |  |  | <0.001 |  | <0.001 |
| Undetectable | 279 (72.3) | 80 (55.2) |  | 83 (52.9) |  |
| Low | 68 (17.6) | 40 (27.6) |  | 48 (30.6) |  |
| High | 39 (10.1) | 25 (17.2) |  | 26 (16.56) |  |

<sup>a</sup> p-values obtained by fisher's exact test for categorial variables and k-sample equality of median test for continuous variables. P-values are comparison between participants to the sub-cohorts and non-participants.
Abbreviations: HIV, human immunodeficiency virus; gp120, glycoprotein 120; IQR, interquartile range; ARV, antiretrovirals; n, number of individuals; SD, standard deviation

### Measuring soluble gp120 in plasma from PLWH with undetectable viremia

Based on a previous iteration [7], we optimized an ELISA to measure sgp120 in plasma (Figure S1A). In this assay, we use a gp120 inner domain specific antibody (C11) which targets the highly-conserved N-termini and 8-stranded β-sandwich structure of gp120 that is formed at the late stage of HIV-1 entry [28]. This epitope is therefore buried on the trimeric Env present on virions or infected cells but exposed on sgp120 [16, 28, 29]. The novelty of this assay is the use of the broadly-neutralizing (bNAb) CD4-binding site (CD4BS) N6 antibody as a readout, which does not compete for C11 binding. This Ab was reported to target up to 98% of global HIV-1 isolates [30]. Using our assay, a strong linearity (r=0.9836, p<0.0001) over a 500-fold range was observed between the signal obtained and the quantity of purified recombinant monomeric soluble HIV-1_YU2_ gp120 used (Figure S1B). This assay enabled us to measure sgp120 in plasma from PLWH with undetectable viral loads compared to uninfected individuals (Figure 1A). We further stratified PLWH in three sub-groups based on the positivity threshold established with the uninfected plasmas: 1) undetectable sgp120, 2) sgp120_low_ and 3) sgp120_high_ (Figure 1A). Of the 386 plasma samples analyzed, 72.3% (n = 279) had undetectable levels of sgp120, 17.6% (n = 68) were sgp120_low_ and 10.1% (n = 39) were sgp120_high_ (Table 1).

**Figure 1.**
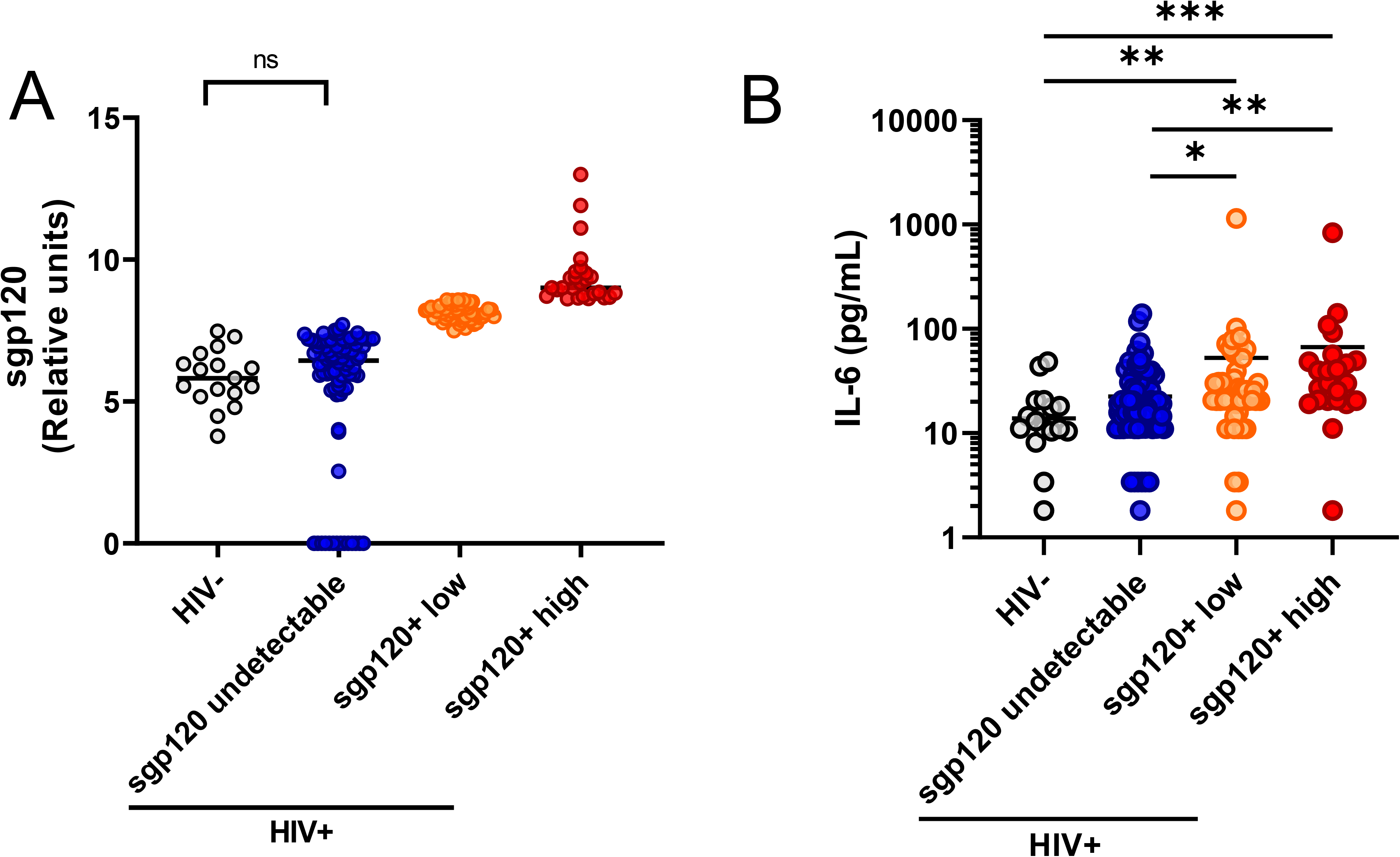
sgp120 detection in PLWH with undetectable viremia is associated with inflammation. Representative stratification of 157 PLWH based on sgp120 levels (panel A), with levels of IL-6 being shown for the respective 157 PLWH (panel B). Statistical analysis was performed using Mann-Whitney *U tests* (panel B). Abbreviations: sgp120, soluble glycoprotein 120; pg, picogram; HIV, human immunodeficiency virus; IL-6, interleukin-6; PLWH, people living with HIV

### Anti-cluster A antibodies levels are associated with immune dysfunction

We have previously reported that the release of sgp120 from infected cells sensitizes uninfected bystander CD4^+^ T cells to ADCC mediated by HIV+ plasma *in vitro*, whether this happens in PLWH remains unclear [15–17]. We therefore measured the levels of anti-cluster A antibodies using an engineered stabilized gp120 inner domain protein (ID2) only exposing the cluster A region [24, 31]. Figure 2 and Table S1, present the associations between anti-cluster A antibodies and absolute CD4 cell counts or the CD4:CD8 ratio (Figure S1). In the total study population, after adjustment for potential confounders (including age, sex, ethnicity, smoking, nadir CD4 and duration of antiretroviral therapy), each 1log_2_ increase in antibody levels was associated with a mean predicted decrease of 15 x10^6^ CD4 cells/ml (95% CI -26 to -3.84, p=0.009) (Figure 2A). Upon stratification of the 386 PLWH by sgp120 levels, we observe that the mean predicted change in CD4 cell counts varies by stratum of sgp120. For those with sgp120 below detection levels, each increase of 1log_2_ in anti-cluster A antibodies is associated with a mean predicted decrease of 18 x10^6^ CD4 cells/ml in absolute CD4 cell count (p=0.008) (Figure 2B), while the mean predicted decline for participants with high levels of sgp120 is 42 x10^6^ CD4 cells/ml (p=0.042) (Figure 2D). However, the p-value for the interaction was 0.11 (Table S1). A similar dynamic is observed with the CD4:CD8 ratio (presented in Figure S2 and Table S1); within the total study population, each 1log_2_ increase in the levels of anti-cluster A antibodies is associated with a mean projected decrease in the CD4:CD8 ratio of -0.06 (95%CI -0.08 to -0.03, p<0.001) (Figure S2A). The magnitude of this association was more pronounced in the subgroup with high levels of sgp120, in which the mean predicted decline in CD4:CD8 ratio was -0.13 (95%CI -0.22 to -0.04, p=0.004) (Figure S2D). However, this difference was not statistically different between the groups (the p-value for interaction between sgp120 levels and anti-cluster A antibodies was 0.21) (Table S1). Of note, no association was observed between anti-cluster A antibodies and CD4 T cell count or CD4:CD8 ratio in individuals presenting low levels of sgp120 (Figure 2C, Figure S2C, Table S1).

**Figure 2.**
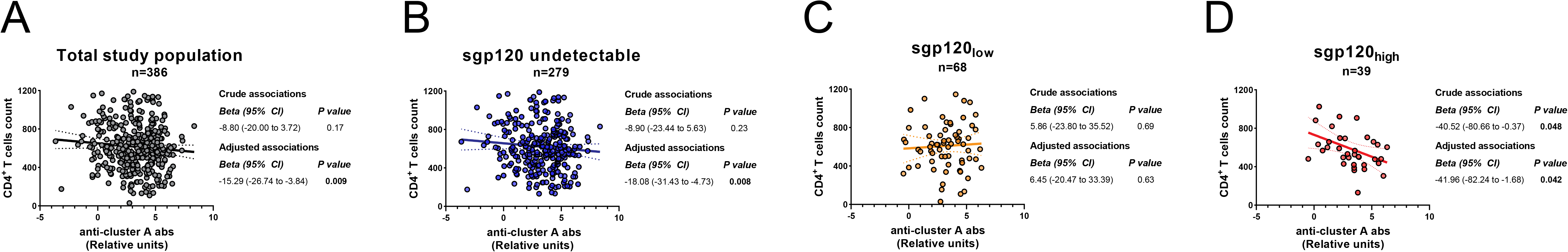
Anti-cluster A antibodies inversely correlate with CD4 counts in PLWH presenting high levels of sgp120. Correlation between the CD4^+^ T cells count with anti-cluster A antibodies upon stratification of 386 PLWH by sgp120 levels. Correlations are depicted for the total study population (Panel A), sgp120 undetectable (Panel B), sgp120_low_ (Panel C) and sgp120_high_ (Panel D). Levels of anti-cluster A antibodies were log_2_ transformed. Uni and multivariable linear regressions were performed, with the beta parameters representing the mean predicted change in absolute CD4 cell counts for each 1log_2_ increase in titers of anti-cluster A abs. Multivariable models are adjusted for age, sex, ethnicity, smoking, duration of antiretroviral therapy and nadir CD4 cell counts. Abbreviations: PLWH, people living with HIV; sgp120, soluble glycoprotein 120; n, numbers of individuals; CI, confidence interval

### Levels of sgp120 are associated with pro-inflammatory markers and modify the associations between them

The HIV-1 gp120 is a pleiotropic molecule beyond its key role in viral entry [4, 7, 14, 15]. The presence of sgp120 was associated with increased levels of pro-inflammatory markers in the plasma of early and acute PLWH [7]. To test whether this observation could be extended to long-term treated PLWH, we performed multiplex measurements of various soluble markers associated with chronic inflammation (Table S4) in a subset of 157 PLWH. Our results show that plasmatic levels of IL-6 are significantly higher in people with low and high levels of sgp120 as compared to the sgp120 undetectable group and uninfected controls (Figure 1B. Figure 3 shows the correlations between all inflammatory biomarkers measured by multiplex platform, as a function of detectable levels of sgp120. Edge bundling correlation plots show the effect of the presence of sgp120, with the undetectable group having weak associations among the parameters measured (Figure 3A), while the presence of low (Figure 3B) and high (Figure 3C) levels of sgp120 intensifies the network of associations, notably between soluble CD14 (sCD14) and the lymphocyte counts, including a strengthened inverse correlation with CD4:CD8 ratios. These results suggest that sgp120 acts as an effect modifier, modulating the associations among the different biomarkers analyzed in this study.

**Figure 3.**
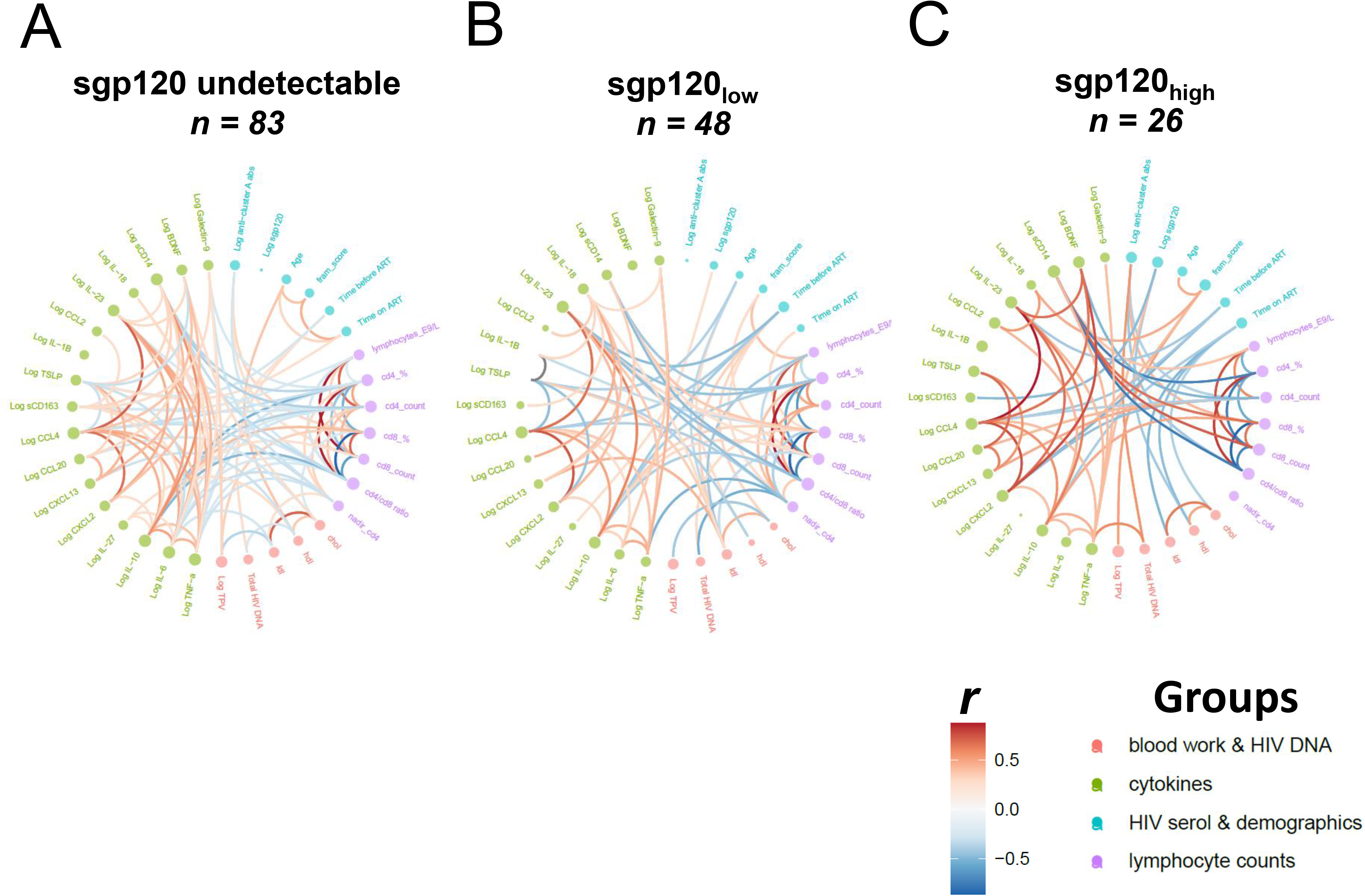
sgp120 act as an “effect modifier’’ of the associations between clinical and laboratory markers. The presence of sgp120 modifies the network of associations between clinical and laboratory parameters in PLWH with undetectable viremia upon stratification by undetectable levels (panel A), low levels (panel B) and high levels (panel C). Spearman Rank correlations were computed and graphed as edge bundling correlation plot using program R. Statistical tests were two-sided and p < 0.05 was considered significant. Edges are only shown if p < 0.05, and nodes are sized according to the connecting edges’ r values. Nodes are color-coded according to the grouping of variables. Abbreviations: sgp120, soluble glycoprotein 120; PLWH, people living with HIV; chol, cholesterol; hdl, high density lipoproteins; ldl, low density lipoproteins; TPV, total atherosclerotic plaque volume; HIV, human immunodeficiency virus; BDNF, brain derived neurotrophic factor; sCD163, soluble CD163; TSLP, thymic stromal lymphopoietin; sCD14, soluble CD14; fram score, Framingham risk score; n, numbers of individuals

### In participants with subclinical cardiovascular disease and detectable sgp120, anti-cluster A antibodies and sgp120 levels are associated with the size of coronary artery plaque

In the subgroup of 145 participants with available cardiovascular imaging, the presence or absence of detectable cardiovascular disease (CVD) (defined as presence of at least one measurable coronary atherosclerotic plaque on CCTA) was not associated with levels of sgp120 (adjusted odds ratio 1.04, 95% CI 0.96 to 1.12) anti-cluster A antibodies (adjusted odds ratio 1.01, 95% CI 0.82 to 1.24) or a score defined by multiplying the levels of sgp120 and anti-cluster A antibodies (adjusted odds ratio 1.03, (95% CI 0.96 to 1.1). However, among the 46 individuals with detectable subclinical CVD and detectable sgp120, we found an association between the size of the coronary artery plaques and the levels of sgp120 (p=0.018), anti-cluster A antibodies (p=0.01) and their combined score (p=0.006) (Figure 4, Table S2).

**Figure 4.**
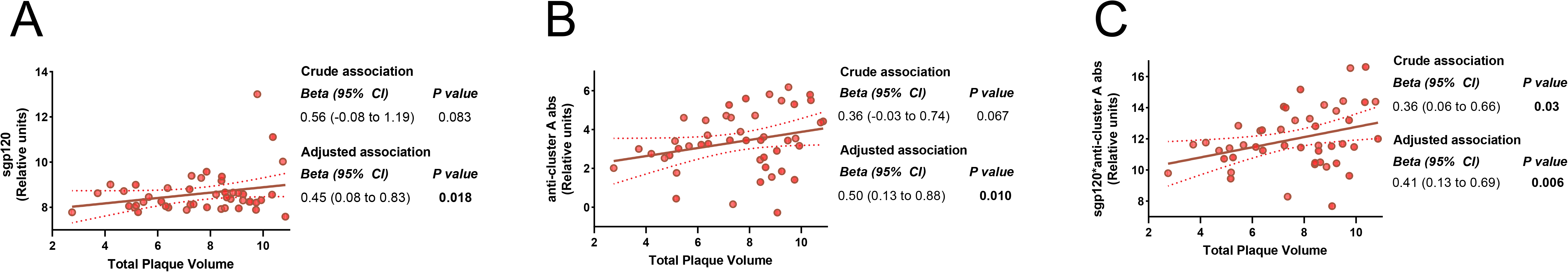
Levels of sgp120 and anti-cluster A abs correlate positively with subclinical cardiovascular disease. Association between the size of coronary artery plaque volume and soluble gp120 levels (panel A), anti-cluster A antibodies (panel B) and the multiplicative combination of both (panel C) in PLWH positive for sgp120 and with detectable subclinical cardiovascular disease. Values for sgp120, anti-cluster A abs and total plaque volume were log_2_ transformed. Uni and multivariable linear regressions were performed, with the beta parameters representing the mean predicted change in log total plaque volume for each 1log_2_ increase in the exposure. Multivariable models are adjusted for age, sex, smoking, low/high density lipoproteins, diabetes and hypertension. Abbreviations: PLWH, people living with HIV; sgp120, soluble glycoprotein 120; n, numbers of individuals; CI, confidence interval

## DISCUSSION

In this cross-sectional study, nested within a large observational cohort, we explored whether sgp120 could be associated with chronic inflammation and cardiovascular disease in long-term treated PLWH. We demonstrated that sgp120 was detectable in the plasma of 107/386 (28%) participants with undetectable viremia and that these participants had more pronounced immune dysfunction associated with the presence of anti-cluster A antibodies, as well as distinct soluble inflammatory marker profiles. Additionally, for those with detectable coronary atherosclerosis, sgp120 and anti-cluster A antibodies were associated with a significant increase in the size of coronary atherosclerotic plaques. Taken together, these results suggest that gp120 might represent a targetable pathway to modulate immune dysfunction, chronic inflammation, and cardiovascular disease risk in PLWH.

We optimized an ELISA assay to detect sgp120 using C11, a monoclonal antibody recognizing a highly-conserved gp120 region that is buried at the Env trimer interface and gets exposed upon dissociation of the gp120 with gp41 [28], thus recognizing sgp120. We used the N6 bNAb known to recognize 98% of global HIV-1 strains [30] as a readout. The use of a detection antibody targeting the CD4BS enables the detection of sgp120 which has the potential of binding the CD4 receptor, whose interaction has been associated with immune activation and inflammation *in vitro* [4, 14, 15]. While sgp120 alone was not significantly associated with most clinical parameters measured, stratification of 386 PLWH based on sgp120 levels unveiled a striking inverse correlation between the levels of anti-cluster A antibodies with CD4 count and CD4:CD8 ratio. These associations remained after adjustment for potential confounders (including age, sex, ethnicity, smoking, nadir CD4 and duration of antiretroviral therapy). This *in vivo* association is consistent with previous well-established *in vitro* evidence showing that binding of sgp120 to uninfected bystander CD4^+^ T cells lead to their cell death via ADCC upon recognition by HIV+ plasma [15–17].

We also observed a significant increase in IL-6 in individuals presenting detectable levels of sgp120. On one hand, plasmatic level of IL-6 has been reported to be independently associated with frailty [32] as well as serious non-AIDS conditions or death among PLWH with undetectable viremia [33]. On the other hand, *in vitro* studies demonstrated that sgp120 binding to the CD4 receptor or CCR5/CXCR4 coreceptors on immune cells induces the liberation of inflammatory markers [4, 14, 15]. All together, these data suggest that sgp120 could contribute to the development of comorbidities by inducing proinflammatory cytokines *in vivo*.

Anti-cluster A antibodies, present in all PLWH [34, 35], and sgp120 were shown to eliminate uninfected bystander CD4^+^ T cells by ADCC *in vitro*. Here we extend these results by showing an association with CD4^+^ T cell levels in a subset of PLWH (those presenting high levels of sgp120). Further, the combination of these two parameters was associated with the size of atherosclerotic plaques, which defines subclinical cardiovascular disease [20, 23], and this remained after adjusting for potential confounders factors. This suggests that the levels of sgp120 and anti-cluster A antibodies could contribute to the development of CVD, which could be linked to depletion of uninfected bystander CD4^+^ cells, as it was shown that a low CD4:CD8 ratio was predictive of the presence of CVD in PLWH undertaking ART [36, 37].

Notably, we propose that sgp120 acts as an ‘’effect modifier’’, potentially driving immune dysfunction, chronic inflammation, and the development of premature comorbidities. Thus, small molecules targeting gp120 could potentially ‘’detoxify’’ its immunomodulatory effect and alleviate the development of premature comorbidities. One such molecule is the recently FDA-approved small-molecule gp120 inhibitor fostemsavir, a pro-drug of temsavir (TMR), a novel attachment inhibitor that can prevent Env-CD4 interaction (BMS-663068; GSK3684934, Rukobia) and is currently used in combination with other ART for adults with multidrug-resistant HIV-1 [38, 39]. Intriguingly, many PLWH receiving fostemsavir exhibit clinical benefits that go beyond its capacity to decrease viral loads below detection levels [38, 39]. Our group has recently shown that TMR reduces the release of gp120, and that shed gp120 produced from TMR-treated cells loses its capacity to interact with uninfected bystander CD4^+^ T cells, protecting these cells from ADCC responses and sgp120-mediated cytokine burst in human PBMCs [15]. Whether the use of TMR in treated PLWH presenting high levels of sgp120 could help alleviate sgp120-mediated chronic inflammation and immune dysfunction is yet to be shown.

The source of circulating sgp120 in long-term ART treated PLWH with undetectable viral loads is not well understood. Although ART suppresses viral replication, it does not prevent proviral gene expression from the viral reservoir [40]. As such, a fraction of cells harboring intact or defective proviruses that persist during ART have been found to be transcriptionally competent, leading to the production of viral proteins and activation of the immune system [41, 42]. Interestingly, defective proviruses able to produce *env* transcripts have been found in a subset of PLWH on ART [43], raising the possibility that translation-competent proviruses could contribute to the production and release of sgp120 even under ART. Accordingly, we did observe a significant association between the levels of HIV DNA, with the combination of sgp120 and anti-cluster A antibodies (Figure 3, Table S3), although sgp120 alone was not significantly associated. Whether this is due to the limited number of individuals with detectable sgp120 in our cohort remains to be determined. Alternatively, the lack of association between sgp120 alone and the reservoir might be due to the fact that measures of HIV reservoir were performed in the blood and not in the tissues, which may be a preferential site for persistence of the active reservoir [44]. Finally, it has been demonstrated that the size of the viral reservoir in PLWH on ART was associated with subclinical CVD [26, 45], whether this association is linked to the persistence of sgp120 in those individuals remains to be determined. Future studies are needed to elucidate the mechanism by which sgp120 persists over time and whether it may represent a potential therapeutic target to reduce the risk of early onset comorbidity for PLWH.

Limitations of our study include its cross-sectional design, the reliance on subgroups in whom data was available for cardiovascular disease assessment, and limited power for analysis of the sgp120 strata. While reverse causality is unlikely (cardiovascular disease could not cause the presence of sgp120), a causal relationship cannot be inferred from this design, and longitudinal assessment of sgp120, anti-cluster A antibodies and coronary artery disease assessment is needed, as well as replication of findings in independent populations.

In conclusion, in this cross-sectional study, nested within a large cohort study of adults aging with HIV infection with undetectable viral load, we demonstrate that sgp120 is detectable in about one third of participants, and that along with levels of anti-cluster A antibodies, it modulates immune and inflammation profiles. This may represent a novel treatment target for PLWH, to address risk of inflammation-related comorbidities.

## NOTES

### Author contributions

Conceptualization, M.B., J.R., W.M., M.D., and A.F.; methodology, M.B., J.R., and A.F.; investigation, M.B., J.R., C.B., C.C.-L., G.G.-L., M.F., M.M.-P., C.G., I.T., R.F., M.M.-V., and J.P.; resources, M.B., J.R., C.B., W.D.T., G.G.-L., M.F., N.C., M.P., M.D., and A.F.; formal analysis, M.B and M.D.; visualization, M.B, R.D., M.D. and A.F.; supervision, M.B., D.E.K., F.M., R.D., N.C., P.B., M.-J.B., M.P., M.D., and A.F.; funding acquisition, M.D. and A.F.; writing – original draft, M.B., M.D. and A.F.; writing – review & editing, M.B., J.R., C.B., W.D.T., C.C.-L., G.G.-L., M.F., M.M.-P., C.G., I.T., R.F., M.M.-V., J.P., A.C., D.E.K., F.M., R.D., N.C., P.B., C.T., J.-G.B., B.T., S.T., M.P., M.D., and A.F

## Acknowledgments.

The authors thank the CRCHUM BSL3 platform. This study was supported by grants from the National Institutes of Health: R01 AI148379), R01 AI150322 to A.F; R01 AI129769 to A.F and M.P; and R01 AI116274 to MP. It was also supported by a FRQS AIDS and Infectious Diseases Network grant to J.R., M.D. and A.F. This work was also partially supported by a CIHR foundation grant #352417 and a CIHR Team grant #422148 to A.F. The study was also supported by Canada Foundation for Innovation grant #41027 to D.E.K., N.C., and A.F. The Canadian HIV and Aging cohort Study is supported by CIHR team grant on HIV and healthy living (HAL398643 CIHR-IRSC:0635001811) and receives in kind support from the CIHR HIV Clinical Trial Network (CTN-272). This study was also supported by a pilot study grant from the CIHR HIV Clinical Trial Network (CTNPT 052). A.F. is the recipient of a Canada Research Chair on Retroviral Entry #RCHS0235 950-232424. CT is the Pfizer/Université de Montreal chair on HIV translational research. M.B. was supported by a CIHR fellowship. M.D. is supported by a clinician-researcher salary award from the *Fonds de recherche du Québec-Santé*. Supplemental Figure 1 was prepared using illustrations from BioRender. The funders had no role in study design, data collection and analysis, decision to publish, or preparation of the manuscript.

## Disclaimer

The views expressed in this manuscript are those of the authors and do not reflect the official policy or position of the Uniformed Services University, US Army, the Department of Defense, or the US Government.

## Data availability

Data and reagents are available upon request.

---

**Supplemental Figure 1.**
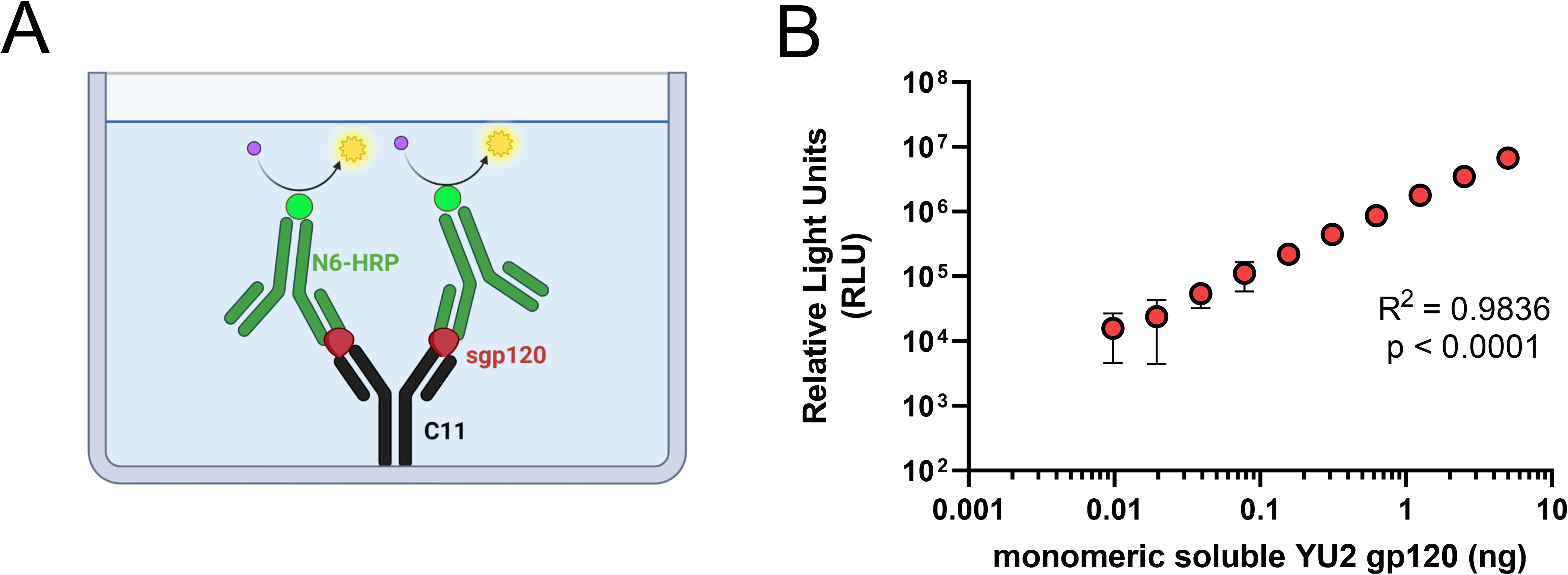
Soluble gp120 ELISA. Schematic of the sgp120 ELISA assay (panel A) and standard curve obtained with reference recombinant sgp120 (n=4) (panel B). Statistical analysis was performed using simple linear regression (panel B). Abbreviations: HRP, horse radish peroxidase; gp120, glycoprotein 120; ng, nanogram

**Supplemental Figure 2.**
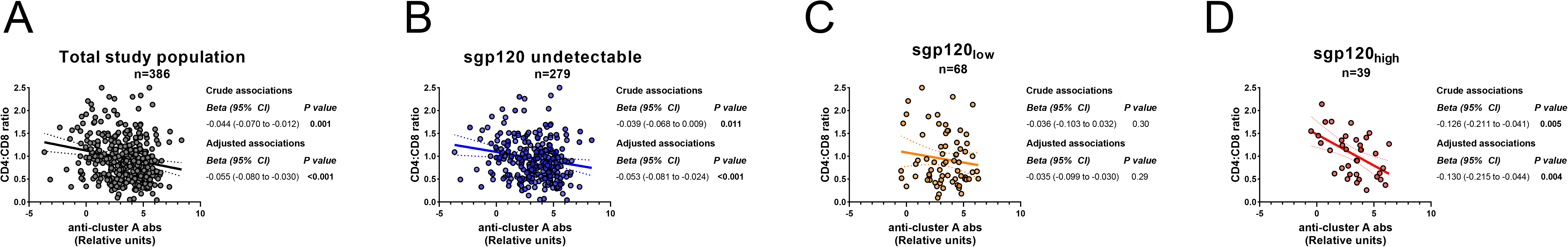
Anti-cluster A antibodies inversely correlate with CD4:CD8 ratio in PLWH presenting high levels of sgp120. Correlation between the CD4:CD8 ratio with anti-cluster A abs upon stratification of 386 PLWH by sgp120 levels. Correlations are depicted for the total study population (Panel A), sgp120 undetectable (Panel B), sgp120_low_ (Panel C) and sgp120_high_ (Panel D). Levels of anti-cluster A antibodies were log_2_ transformed. Uni and multivariable linear regressions were performed, with the beta parameters representing the mean predicted change in CD4:CD8 ratio for each 1log_2_ increase in titers of anti-cluster A abs. Multivariable models are adjusted for age, sex, ethnicity, smoking, duration of antiretroviral therapy and nadir CD4 cell counts. Abbreviations: PLWH, people living with HIV; sgp120, soluble glycoprotein 120; n, numbers of individuals; CI, confidence interval

**Table S1.**
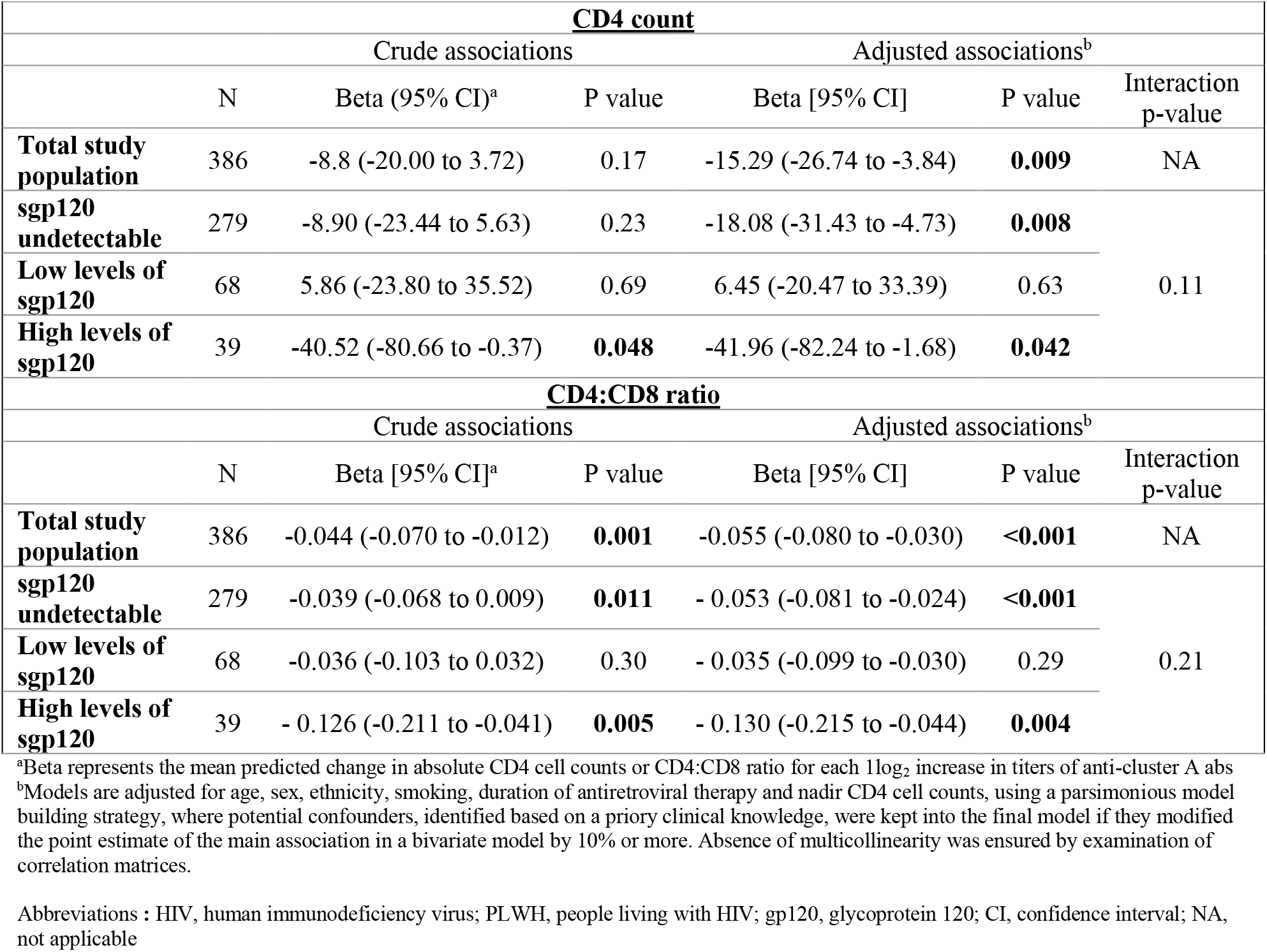
Association between the levels of anti-cluster A antibodies and absolute CD4 count or CD4:CD8 ratio, stratified by levels of soluble gp120 in 386 PLWH with undetectable HIV viremia.

**Table S2.**
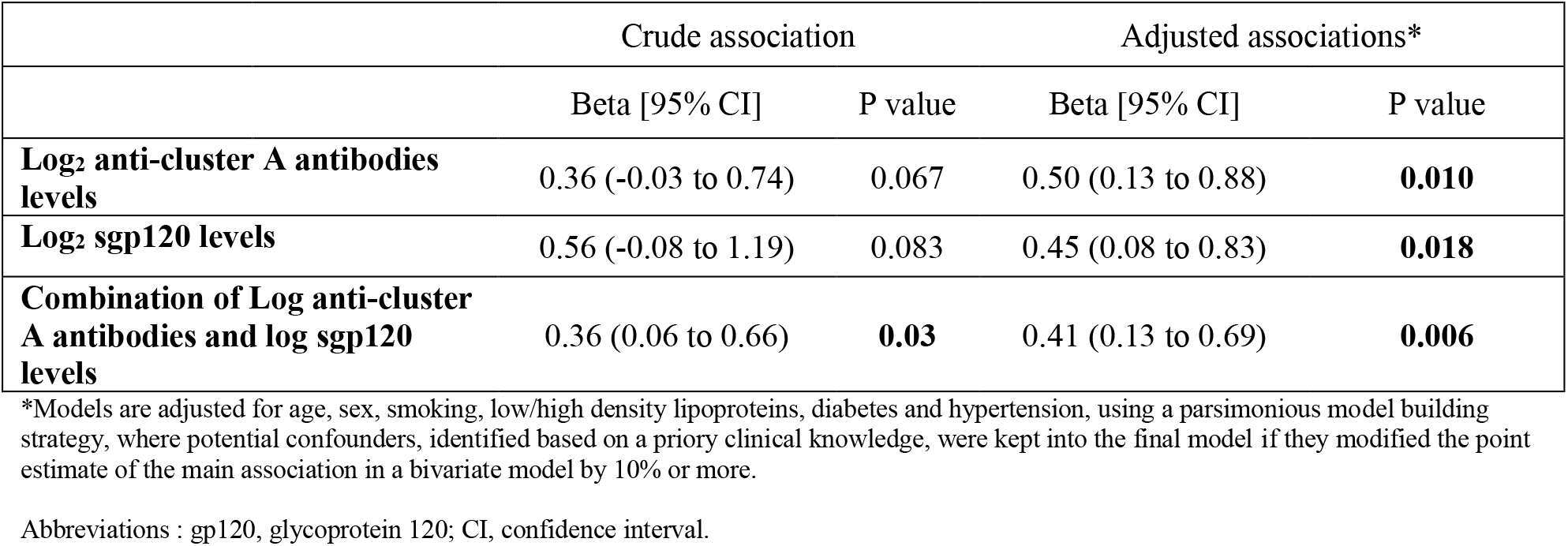
Association between anti-cluster A antibodies, sgp120 levels and their combination with the size of coronary artery plaque in 46 participants with both cardiovascular disease and detectable gp120.

**Table S3.** Associations* between sgp120 and anti-cluster A antibodies with clinical and laboratory markers in 157 PLWH with multiplex measurements.

|  |  |  |  |
| --- | --- | --- | --- |
| **<u>Anti-cluster A antibodies</u>** |  |  |  |
|  | sgp120 undetectable | sgp120<sub>low</sub> | sgp120<sub>high</sub> |
| **%CD8** | S<sub>Rho</sub> = 0.311; p= 0.004 | n.s | S<sub>Rho</sub> = 0.447; p=0.022 |
| **CD8 count** | S<sub>Rho</sub> = 0.256; p=0.021 | n.s | n.s |
| **CD4:CD8 ratio** | S<sub>Rho</sub> = -0.272; p=0.013 | n.s | S<sub>Rho</sub> = -0.449; p=0.021 |
| **CD4 count** | n.s | n.s | S<sub>Rho</sub> = -0.396; p=0.045 |
| **Total HIV DNA** | n.s | n.s | S<sub>Rho</sub> = 0.577; p=0.008 |
| **<u>soluble gp120</u>** |  |  |  |
|  | sgp120 undetectable | sgp120<sub>low</sub> | sgp120<sub>high</sub> |
| **High density lipoproteins** | n.s | n.s | S<sub>Rho</sub> = -0.436; p=0.026 |
| **TNFα** | n.s | n.s | S<sub>Rho</sub> = 0.410; p=0.038 |
| **sCD163** | n.s | n.s | S<sub>Rho</sub> = -0.510; p=0.008 |

\*Spearman Rank correlations.
Abbreviations : n.s, not significant; sgp120, soluble gp120

**Table S4.** List of analytes measured in plasma by beads arrays.

| **Table S4 - List of analytes measured in plasma by beads arrays** |  |
| --- | --- |
| **Analytes** | **Bead Region** |
| **CCL2/JE/MCP-1<sup>a</sup>** | 62 |
| **CCL20/MIP-3α<sup>a</sup>** | 33 |
| **CXCL2/Gro-β/MIP-2/CINC-3<sup>a</sup>** | 27 |
| **IL-1β<sup>a</sup>** | 57 |
| **IL-10<sup>a</sup>** | 22 |
| **IL-23<sup>a</sup>** | 76 |
| **TNFα<sup>a</sup>** | 12 |
| **CCL4/MIP-1β<sup>a</sup>** | 37 |
| **Soluble CD163 (sCD163)<sup>a</sup>** | 28 |
| **CXCL13/BLC/BCA-1<sup>a</sup>** | 14 |
| **IL-6<sup>a</sup>** | 13 |
| **IL-18/IL-1F4<sup>a</sup>** | 78 |
| **IL-27<sup>a</sup>** | 25 |
| **TLSP<sup>a</sup>** | 48 |
| **BDNF<sup>b</sup>** | 15 |
| **Galectin-9<sup>b</sup>** | 18 |
| **Soluble CD14 (sCD14)<sup>c</sup>** | 29 |

Human Magnetic Luminex ® Assays, from R&D Systems (Bio-Techne) Premixed Multiplex Kit Catalog Numbers : <sup>a</sup>LXSAHM-14, <sup>b</sup>LXSAHM-02, <sup>c</sup>LXSAHM-01 Kit Lot Number : <sup>a</sup>C0010938, <sup>b</sup>L150692, <sup>c</sup>L148511

